# Comprehensive Systematic Review to Identify putative COVID-19 Treatments: Roles for Immunomodulator and Antiviral Treatments: The University of Birmingham 622 COVID-19 taskforce

**DOI:** 10.1101/2020.08.13.20174060

**Authors:** Thomas Hill, Mark Baker, Lawrence Isherwood, Lennard YW Lee

**Affiliations:** Department of Oncology, Queen Elizabeth Hospital Birmingham, Mindelsohn Way, Edgbaston B15 2GW, United Kingdom; Birmingham Medical School, College of Medical and Dental Sciences, University of Birmingham, Edgbaston, Birmingham B15 2TT; Institute of Cancer and Genomic Sciences, University of Birmingham, Edgbaston, Birmingham B15 2TT

**Keywords:** Systematic review, COVID-19, SARS-CoV-2, treatment, trial, antiviral, immunomodulators

## Abstract

**Objectives:** To identify putative COVID-19 treatments and identify the roles of immunomodulators and antivirals in disease management.

**Design:** Systematic review.

**Data sources:** PubMed, bioRxiv.org and medRxiv.org were searched for studies suggestive of effective treatments for COVID-19. Additional studies were identified via a snowballing method applied to the references of retrieved papers as well as a subsequent targeted search for drug names.

**Review methods:** Inclusion criteria included any case series or randomised control trials in any language that were published from 18th December 2019 to 18th April 2020 and described COVID-19 treatment. Of an initial 2140 studies identified from the initial search, 29 studies were found to meet the inclusion criteria and included in this comprehensive systematic review.

**Results:** 19 studies of antiviral treatments for COVID-19 have been reported and seven studies for immunomodulatory treatments. Six randomised controlled trials have been published with one positive trial for Hydroxychloroquine. This small study consisted of 31 patients though subsequent studies showed contradictory findings. All the remaining studies were observational studies, retrospective case reviews or non-randomised trials and these results are difficult to interpret due to methodological issues.

**Conclusions:** To date, an impressive number of studies have been performed in a short space of time, indicative of a resilient clinical trials infrastructure. However, there is a lack of high quality evidence to support any novel treatments for COVID-19 to be incorporated into the current standard of care. The majority of the studies of treatments for COVID-19 could only be found in pre-print servers. Future clinical reviews should therefore be Comprehensive Systematic Reviews involving pre-print studies to prevent potential unnecessary replications of clinical studies.

## Introduction

SARS-CoV-2 is a betacoronavirus of the *Coronaviridae* family; a group of enveloped positive single strand RNA viruses. The virus causes acute respiratory syndrome and was first identified in Hubei province, China in December 2019. Clinically, the presentation of SARS-CoV-2, COVID-19 varies from none or minor ‘common cold’ symptoms, to acute respiratory distress syndrome (ARDS), resulting in severely impaired respiratory function. The highly infectious nature and potential severity of the pathogenicity of COVID-19 has put significant burden on healthcare, social and economic resources. Mortality increases with age, with the highest mortality among people over 80 years of age with a case fatality rate of 21.9% (1). The standard of care treatment of COVID-19 is largely supportive, consisting of maintaining adequate oxygenation, cardiovascular perfusion and treatment of concurrent infection.

The exact mechanisms by which SARS-CoV-2 causes morbidity and mortality are poorly described. Autopsy studies have identified diffuse alveolar damage. On one hand, this might suggest a direct viral cytopathic effect on alveolar cells (2). Conversely, pneumocyte damage may also arise from ARDS, a form of rapid widespread inflammation in the lung, which arises as a consequence of activation of tissue resident macrophages, resulting in chemokine secretion leading to tissue ingress of peripheral immune cells including neutrophils and lymphocytes and further organ damage (2). There may also be a temporal dimension to the disease course of COVID-19, which may progress through different phases. It is hypothesised the initial phases and symptoms are predominantly driven by viral replication (early infection viral phase), with a late phase, and morbidity/mortality, driven by the host immune response.

The global pharmaceutical COVID-19 drug development pipeline has been developed de novo since December 2019. Broadly, two forms of treatments are under investigation. Firstly, antiviral therapy where the intention is to limit and contain viral replication, and secondly, immunomodulatory therapy with the aim to control the hyperinflammatory immune response.

In this review, we have performed a systematic review of all treatments for COVID-19 up to April 18^th^ 2020 and described the potential utility of antiviral and immunomodulatory strategies. This comprehensive systematic review includes all published trials, but also includes pre-print studies which are significantly more numerous and not reviewed in any previous systematic review.

## Methods

### Search Strategy

A systematic review of PubMed, bioRxiv.org and medRxiv.org was performed to find original research articles providing information of interventional treatments against COVID-19. The search strategy was based on the following keywords, “COVID-19”, “coronavirus”, “SARS-CoV-2” and “treatment” (MeSH search terms of (((‘COVID-19’) OR ‘Coronavirus’) OR ‘SARS-CoV-2’) AND ‘Treatment’). To expand the search, a snowballing method was applied to the references of retrieved papers with a subsequent targeted search for drug names. Research abstracts were independently reviewed by two authors to select studies that met our inclusion criteria before the full-text review of selected studies. Discrepancies and doubts of the relevance of the sources were solved by consensus with two or more authors. This systematic review was performed according to PRISMA guidelines (3).

### Study Selection

Clinical studies were included in this study if they were, i) case series or RCTs, ii) from 18^th^ of December 2019 to 18^th^ of April 2020, iii) in any language and iv) describing COVID-19 treatment. Studies were excluded if, i) they reported on dietary modification including traditional herbal medications, ii) retrospective review of existing anti-hypertensives, iii) systematic reviews, iv) reporting on different forms of oxygen/ventilation, v) case reports, or vi) any commentaries or reviews.

The data and clinical findings were identified and entered into a pre-defined data extraction form, which was filled in by two reviewers. Differences were resolved by consensus. The following data were extracted; title, first author name, date of publication, publication journal, country, treatment, treatment dosage, trial design, number of patients, participant demographics, setting of treatment, outcome data, adverse events and control group.

## Results

### Search Results

A search of PubMed, bioRxiv, medRxiv and a targeted search initially yielded a total of 2147 articles related to the treatment of COVID-19 (Figure 1). In total 204 articles were reviewed in detail and 29 articles were found to be relevant to this comprehensive systematic review.

**Figure 1.**
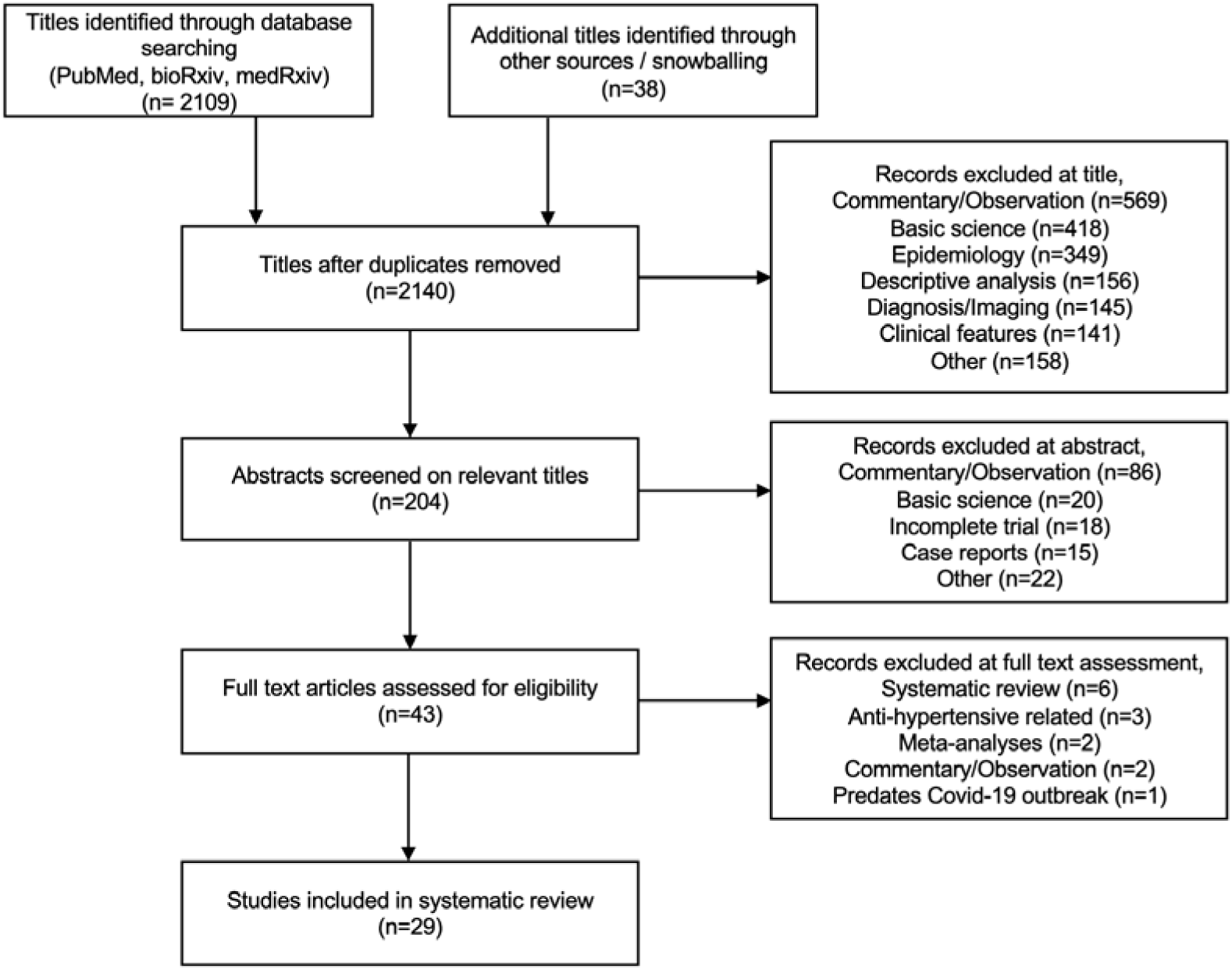
PRISMA flow diagram detailing comprehensive systematic review strategy

**Figure 2:**
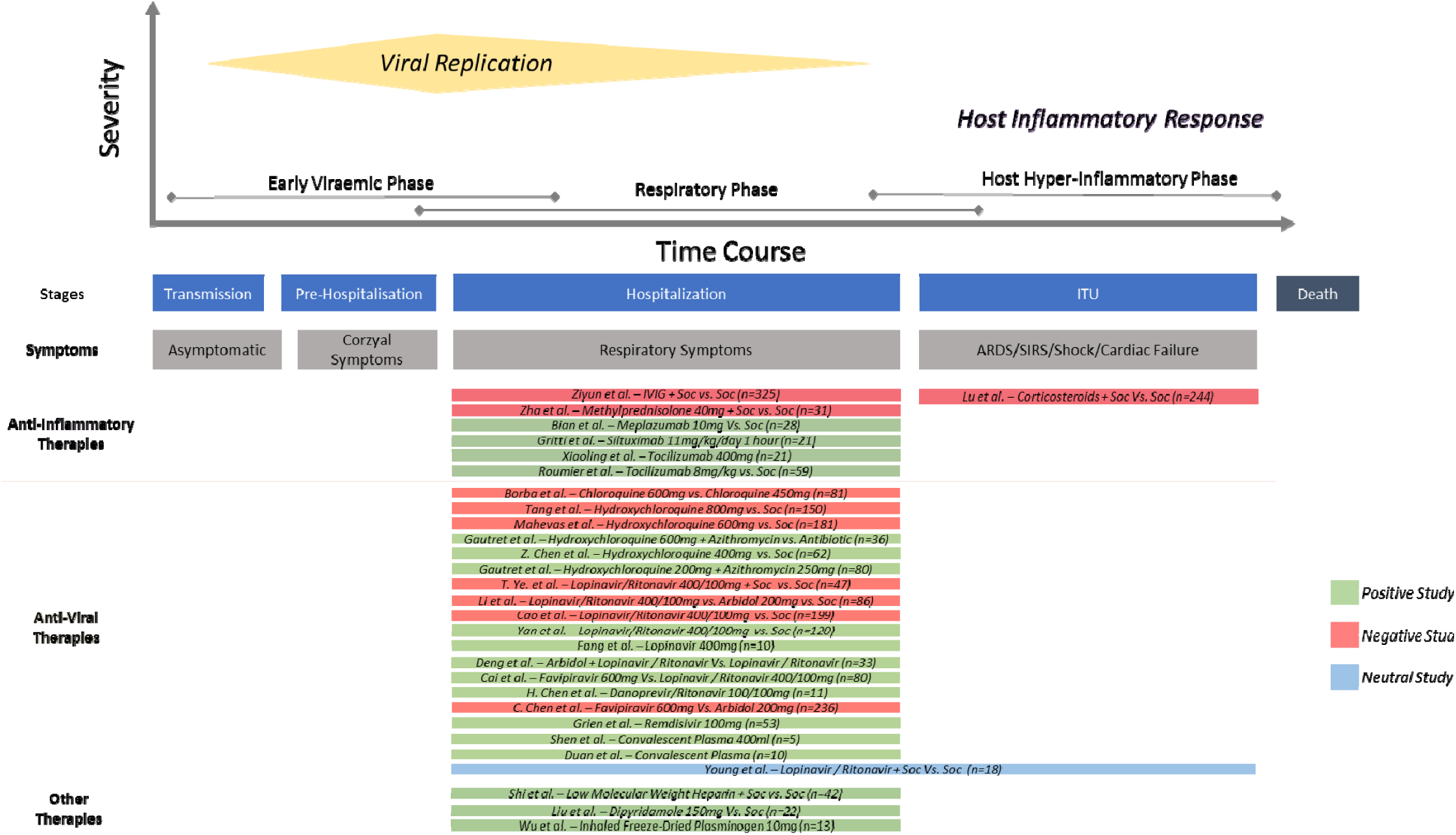
Time course of SARS-CoV-2 infection and timing and results of clinical studies included in this Comprehensive Systematic Review

In total, 13 of these reviews were published in journals, but a significant number were from a pre-print journal (n= 16, 55.2%). In total, there were six RCTs, two non-randomised controlled trials, 10 observational studies, and 11 retrospective case reviews. The total number of patients exposed to these experimental drugs was 2,304, with 56.16% being male (one study did not report gender). The majority of studies were from China (n=21, 72.4%) and Europe (n=5, 17.2%). Overall, a positive trial outcome was reported in 18 studies (62.0%).

The clinical settings for these studies were not well defined. The majority of studies (93.1%, n=27) investigated patients in a hospital setting. Although nine studies were conducted on patients described as severely or critically ill, only one study explicitly stated that it occurred in an intensive therapy unit (ITU) setting. There were no studies performed in an outpatient setting.

The efficacy/outcome measures were extremely heterogeneous and many studies lacked statistical analysis or significance. In terms of reported endpoints of the study, the majority were based on clinical assessment (n=12, 41.4%), some used viral markers (n=7, 24.1%) and a minority used clinical management decisions, such as decision for ITU admission or change in ventilation (n=5, 17.2%). Only two trials used survival as their end point. A significant proportion of studies did not report on adverse events (n=8, 27.5%).

There were 19 studies reporting on antiviral treatment for patients with COVID-19, seven studies investigated immunomodulatory treatment, and three studies looked at Dipyridamole, plasminogen, and low molecular weight heparin. There were six RCTs for antiviral treatments, but no RCTs for an immunomodulatory COVID-19 strategy.

### Antiviral Treatment

Several different types of antiviral treatments for COVID-19 have been analysed to date, three targeting viral entry (Arbidol, Hydroxychloroquine and Chloroquine), two targeting viral polymerases (Favipiravir, Remdesivir) and three protease inhibitors (Lopinavir, Danoprevir, Ritonavir) and one of convalescent plasma.

The largest study of antiviral treatment for COVID-19 to date was for Favipiravir versus a proposed standard of care, Arbidol (4). This was a prospective randomized trial in 236 patients. This study of Favipiravir did not show any evidence of improvement of clinical recovery at day seven compared to Arbidol. It is also notable that there was also no evidence that would suggest Arbidol could be used as an antiviral treatment for COVID-19.

The anti-malarials, Chloroquine and its less toxic derivative Hydroxychloroquine are believed to have antiviral properties. This is mediated through an inhibition of terminal glycosylation of *Angiotensin-converting enzyme 2 (ACE2)*, an enzyme attached to the outer surface (cell membranes) of cells in the lungs (5). However, it is Iikely that anti-inflammatory properties of these drugs also exist (6). The evidence for Chloroquine was from a 2b safety study, however, this was halted early due to significant toxicities. There were two RCTs for the use of Hydroxychloroquine. The first study was performed in a total of 62 patients and found that an unusual outcome measure, ‘time to clinical recovery’ (TTCR), which consists of reduction of body temperature and cough, was significantly improved (7). Unfortunately, the second RCT of Hydroxychloroquine in 150 patients showed no improvement in its primary outcome - 28-day negative conversion rate (8).

Two RCTs have been performed for Lopinavir/Ritonavir; the first involved a total of 86 patients and the second 199 patients. Both studies showed no improvement in any primary or secondary outcome measures with significant toxicities (9) (10).

Finally, there is a retrospective cohort study suggestive that Remdesivir might have efficacy. Remdesivir is a nucleotide analogue prodrug which inserts into viral RNA chains, causing their premature termination (11). A retrospective case review was performed for 53 patients who were treated with Remdesivir. The majority of these patients were receiving invasive ventilation. In this cohort, 47% of patients were discharged and 13% died. However, 60% of patients had an adverse event (12).

### Immunomodulatory Treatment

Two broad immunomodulatory treatment strategies for COVID-19 have been reported. Immunomodulatory treatments were either broad in nature (corticosteroids and intravenous immunoglobulin (IVIG)) or targeted through the use of Siltuximab and Tocilizumab against IL-6 and Meplazumab against CD-147.

The largest study of immunomodulatory treatment for COVID-19 to date is for IVIG versus the standard of care (13). This was a multicentre retrospective cohort study of 325 adults with COVID-19. The study reported that its primary outcome, 28 day and 60 day mortality was not improved with IVIG treatment. However, in subgroup analysis of critical patients, IVIG seemed to reduce the 28 day mortality, reduce patient inflammatory responses and also improved some organ function.

Corticosteroids have well described systemic anti-inflammatory effects, through the up-regulation of anti-inflammatory proteins and down-regulation of pro-inflammatory proteins (14). Two studies report on the role of corticosteroids in COIVD-19 patients; both of which demonstrate corticosteroid therapy have no benefit in viral clearance, symptom resolution or mortality. The larger of the two studies, a retrospective case review of 244 patients suggested that not only did corticosteroids not improve patient’s clinical outcomes, but that a higher steroid dose was significantly associated with elevated mortality risk (15). This study reported that every 10mg increase in Hydrocortisone equivalent dose was associated with an additional 4% mortality risk.

Finally, there were two key studies looking at the impact of targeted immunomodulatory treatment on COVID-19. Siltuximab is a monoclonal antibody that binds to and prevents the action of the pro-inflammatory cytokine IL-6 (16). In 21 hospitalised COVID-19 patients treated on Siltuximab, clinical improvement was observed in 33% of study participants (17). Toculizumab is a monoclonal antibody that binds to IL-6 (18). In an observational study of 30 COVID-19 patients in an intensive care setting, treatment with Toculizumab was found to reduce patient’s mechanical ventilation requirements and need for ITU admission, of which 20% were discharged (19).

**Table 1:**
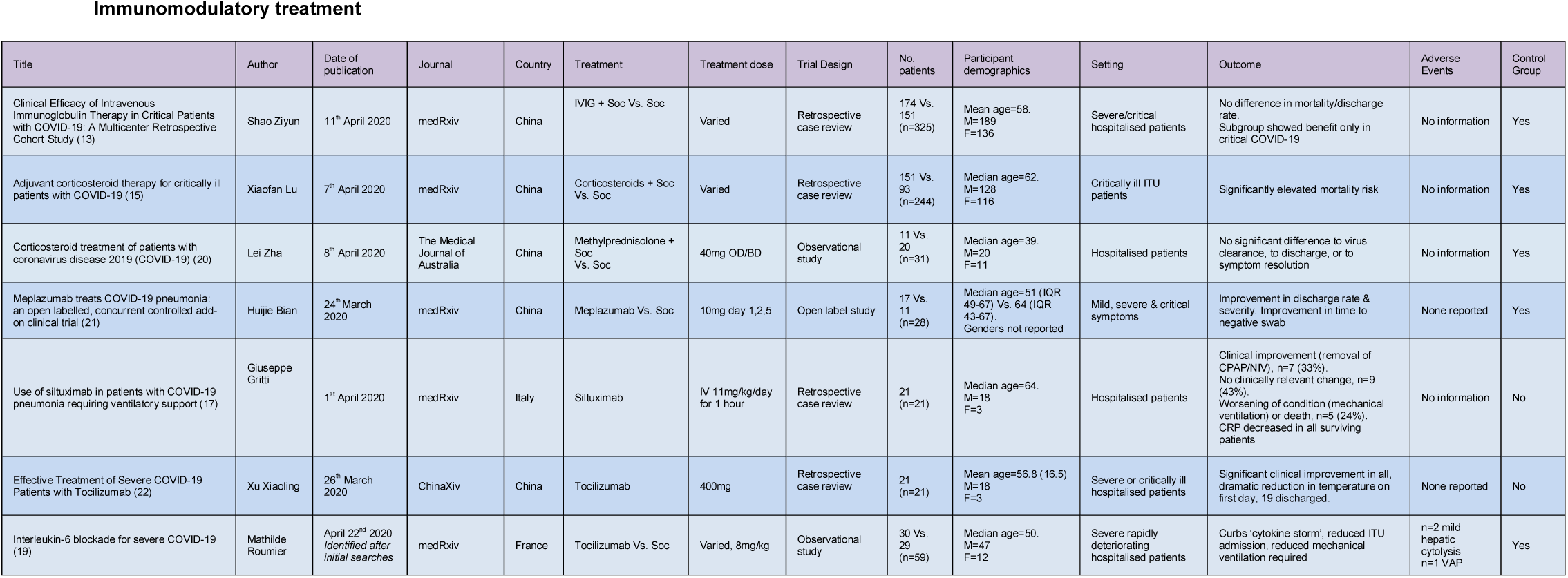
Summary of clinical trials for immunomodulatory treatments against COVID-19.

**Table 2:**
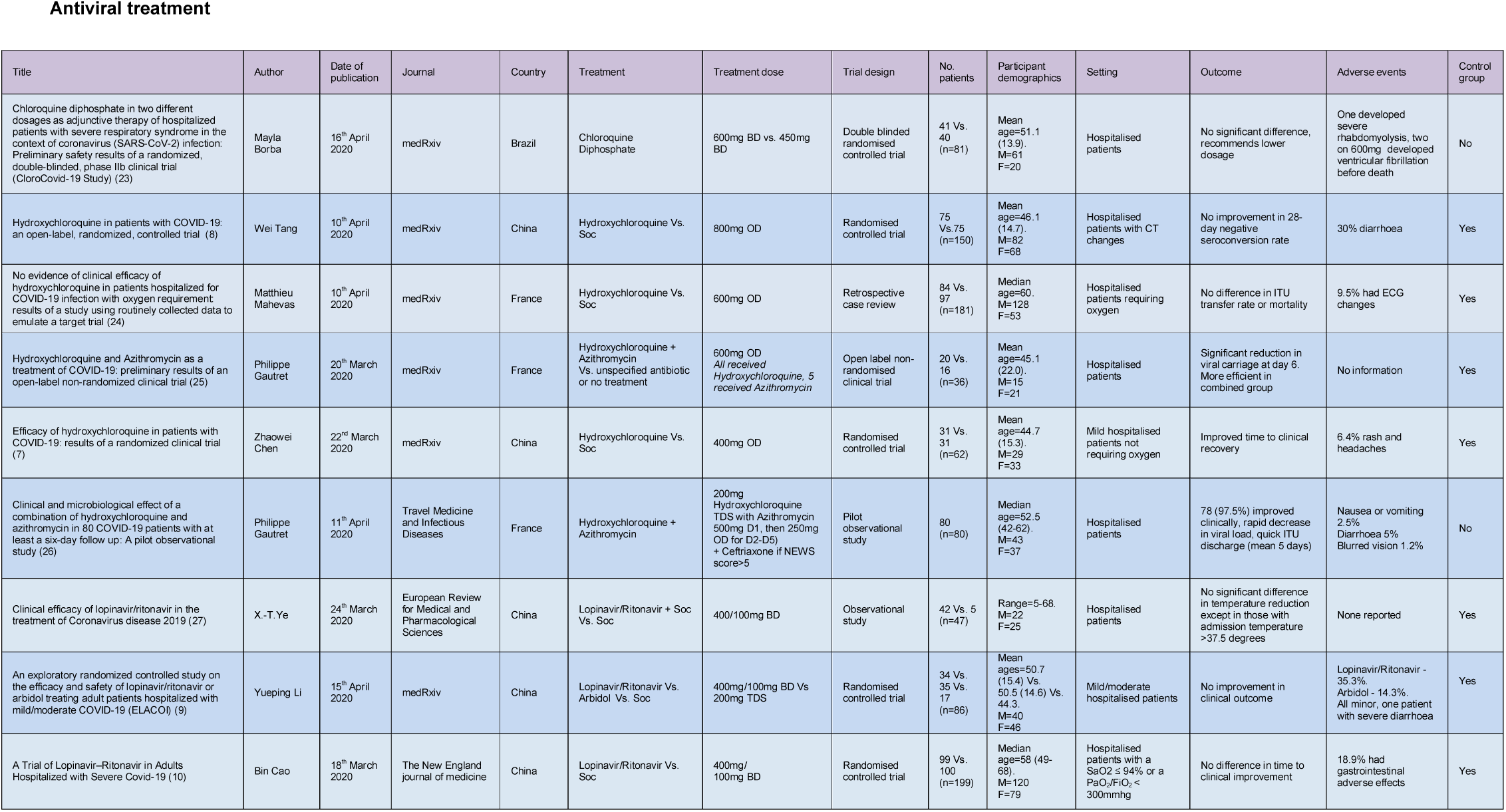
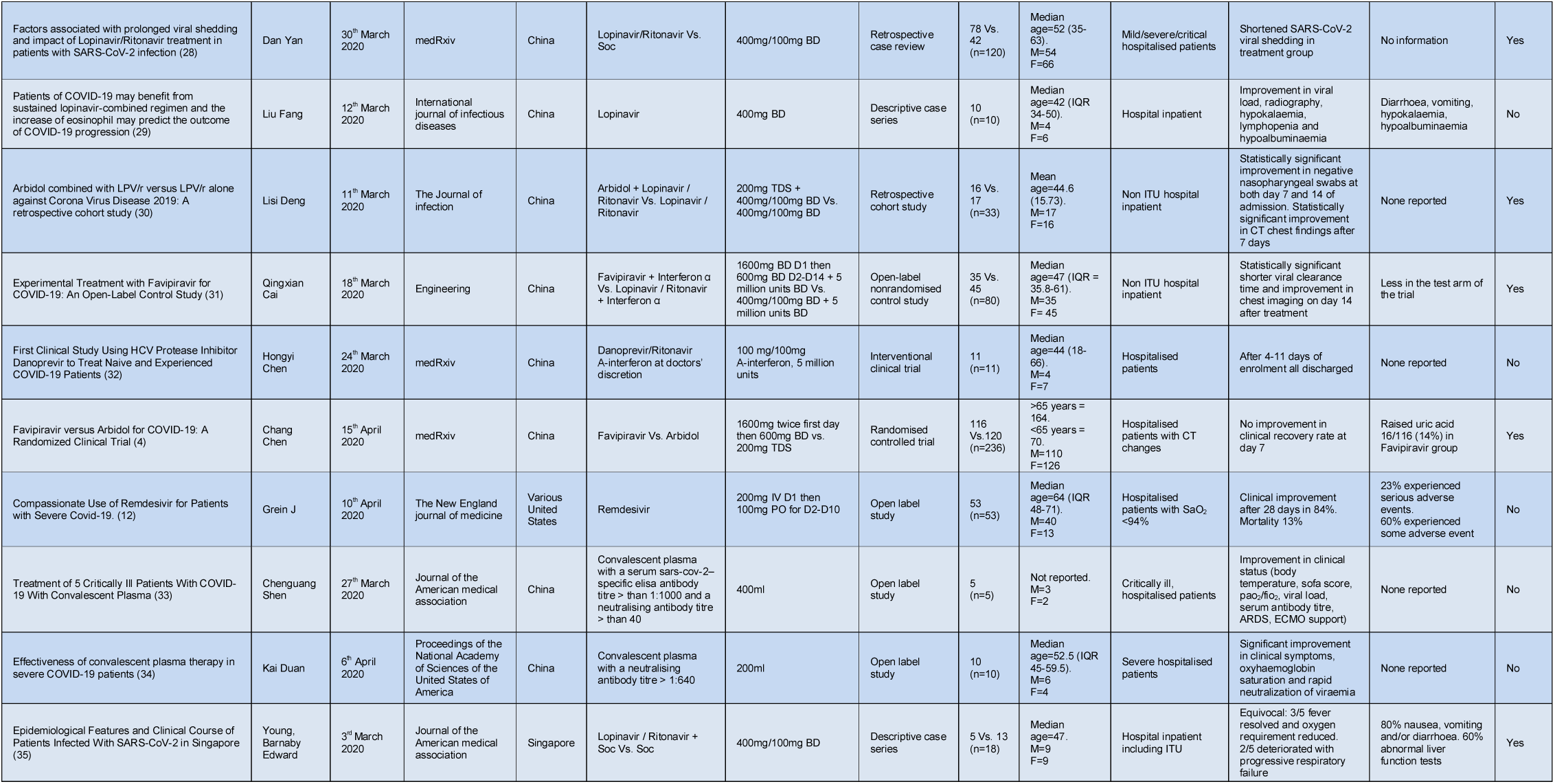
Summary of clinical trials for antiviral treatments against COVID-19.

**Table 3:**
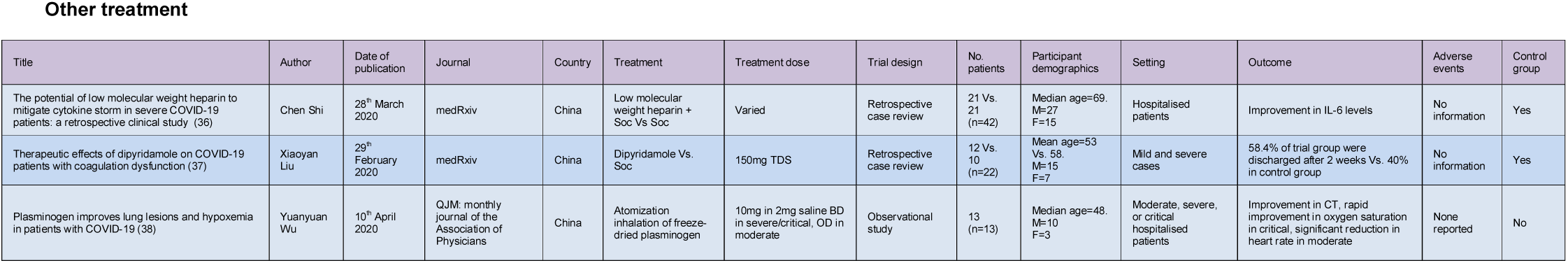
Summary of clinical trials for other treatments against COVID-19.

## Discussions

The rapid global transmission of SARS-CoV-2 has caused significant strain to healthcare systems around the world and is now associated with major morbidity and mortality. The global medical research community has launched a relatively large number of clinical studies in the course of just a few months. The ability to launch these analyses in the middle of a global pandemic, for an essentially unknown disease, is a testament of the robustness of healthcare systems, excellence of individual hospitals/clinicians and is something never seen before in the history of modern medicine.

Unfortunately, this systematic review of clinical trials to date has discovered no high-quality trials identifying drug efficacy against COVID-19. One small RCT of Hydroxychloroquine/Azithromycin was positive but this has not yet been validated and had methodological flaws. However, there are many promising retrospective case studies for antivirals and immunomodulatory treatments, but these remain challenging to interpret due to self-fulfilment bias or trial design. It is also particularly surprising that there are a number of interventions which have negative/no evidence for, which are being tested in very large UK studies. This might be an artefact of a lack of recent clinical reviews or potential oversight of the evidence to date.

Nevertheless, there are a number of valuable insights which may be observed from the studies to date.

Firstly, it would appear that pre-print servers are an extremely good mechanism for disseminating literature/data. With a rapidly ensuing pandemic, the weeks/months required for publication of manuscripts in a peer-reviewed journal may mean that some studies could be needlessly replicated with potential adverse human impact. Whilst not part of the formal process for systematic reviews, the use of pre-print servers would ensure a contemporary review and facilitate a Comprehensive Systematic Review.

Secondly, most of the reported studies investigated hospitalised patients. There is only one study that has emerged out of an intensive care setting. Potentially, this may reflect the relative difficulties of performing studies or case series in ITU. Conversely. More concerningly, it is possible that drug interventions against COVID-19 in an ITU setting may have minimal effect. It might be an unsurmountable challenge for a pharmaceutical intervention to reverse respiratory and multi-organ failure, once a patient enters the hyperimmune phase with an ensuing severe cytokine release syndrome.

Finally, looking to the future, this comprehensive systematic review has identified a number of trial design strategies which would improve future clinical trials. Most of the trials treat a heterogenous patient cohort. As clinical outcomes of patients from COVID-19 vary depending on age, sex and comorbidities, a poorly defined or heterogenous trial inclusion criteria is unlikely to be compatible with efficient trial design. Furthermore, there appear to be a multitude of unvalidated surrogate study endpoints utilised for COVID-19 trials, such as ITU admissions, changes in viral load, seronegativity and cytokine panels. Survival of patients is the definitive end point and one which is rarely utilised but is of great importance. It is therefore of great regret that these trial methodological flaws continue to be repeated in newly launched nationally prioritised studies.

In summary, we have performed a Comprehensive Systematic Review of all pre-print and published articles to date to identify treatments against COVID-19. Unfortunately, there is no high-quality evidence to back any particular intervention, either antiviral or immunomodulatory to form a COVID-19 standard of care. We hope that this work will help ensure the next generation of COVID-19 clinical trials in the UK might have more efficient trial design using better targets and potentially expose less patients to risks beyond COVID-19.

## Data Availability

Data was collected from PubMed, bioRxiv.org and medRxiv.org which are readily available sources of studies.

https://pubmed.ncbi.nlm.nih.gov/

https://www.biorxiv.org/

https://www.medrxiv.org/

## Acknowledgements

The authors thank the doctors, nurses, medical students and healthcare staff working tirelessly on the frontlines of the COVID-19 pandemic at the Queen Elizabeth Hospital Birmingham.

## Author Contributions

TH, MB, LI, LL: Study design, literature review and manuscript drafting. All authors contributed equally.

### Licence statement

The Corresponding Author has the right to grant on behalf of all authors and does grant on behalf of all authors, a worldwide licence to the Publishers and its licensees in perpetuity, in all forms, formats and media (whether known now or created in the future), to i) publish, reproduce, distribute, display and store the Contribution, ii) translate the Contribution into other languages, create adaptations, reprints, include within collections and create summaries, extracts and/or, abstracts of the Contribution and convert or allow conversion into any format including without limitation audio, iii) create any other derivative work(s) based in whole or part on the on the Contribution, iv) to exploit all subsidiary rights to exploit all subsidiary rights that currently exist or as may exist in the future in the Contribution, v) the inclusion of electronic links from the Contribution to third party material where-ever it may be located; and, vi) licence any third party to do any or all of the above. All research articles will be made available on an open access basis.

### Patient and public involvement statement

As we have conducted a systematic review, we have not conducted any primary research with patient involvement. We have no reason to believe that the studies we have included were not carried out *with* patients, carers, or members of the public, rather than on them.

### Dissemination declaration

We do not plan to disseminate results to study participants and patient organisations as it is not possible and not applicable.

